# Translation and Validation of an Epilepsy Screening Instrument in Two Ghanaian Languages

**DOI:** 10.1101/2024.06.01.24306858

**Authors:** Emmanuel Kwame Darkwa, Sabina Asiamah, Elizabeth Awini, Cynthia Sottie, Anthony Godi, John E Williams, Albert Akpalu, J Helen Cross, Josemir W. Sander, Arjune Sen, Charles R. Newton, Anthony Danso-Appiah, Patrick Adjei

## Abstract

**Introduction:** The prevalence of epilepsy in sub-Saharan Africa varies considerably, and the exact estimate in Ghana is unknown, with few data available from peri-urban areas. More community-based studies are required to understand the tangible burden of epilepsy in these areas and the difficulties in healthcare access.

**Objective:** To validate a household survey epilepsy-screening instrument in Shai-Osudoku and Ningo-Prampram District of Greater Accra Region, Ghana.

**Methods:** We developed a 17-item epilepsy screening instrument by modifying validated English language questionnaires. We included questions that could identify convulsive and non-convulsive seizures. Language experts translated and back-translated the survey instrument into the two languages in this region: Asante Twi and Dangme. Cases were people with epilepsy attending healthcare facilities where these languages are used. Controls were unaffected relatives of cases or people attending the healthcare centres for other medical conditions. We matched cases and controls for geographical location and ethnicity. An affirmative response to one of the seventeen questions was deemed as a positive screen.

**Results:** One hundred and Forty Dangme Twi speakers (70 cases and 70 controls) and 100 Twi speakers (50 cases and 50 controls) were recruited. The sensitivity and specificity for Dangme were: Stage 1;100% (95% CI: 88.6, 94.9) and 80% (95% CI: 68.7, 88.6) and Stage 2, 98.6% (95% CI: 92.3, 100.0) and 85.7% (95% CI: 75.3, 92.9). The Dangme version reliably identified epilepsy with positive predictive values of 83.3% (95% CI: 73.6, 90.6) and 87.3% (95% CI: 78.6, 90.6) at stages 1 and 2. The questionnaire excluded epilepsy with negative predictive values of 100% (95% CI: 93.6, 100.0) and 98.4% (95% CI: 91.2, 100.0). For the Twi version, the sensitivity and specificity were: 98% (95% CI: 89.4, 99.9) and 92% (95% CI: 80.8, 97.8) at Stage 1, and for Stage 2, 96% (95% CI: 86.3, 99.5) and 94% (95% CI 83.5, 98.7). The Twi questionnaire reliably specified epilepsy with positive predictive values of 92.5% (95% CI: 81.8%, 97.9) and 94.1% (95% CI: 83.8, 98.8) at stages 1 and 2. It excluded epilepsy with negative predictive values of 97.9% (95% CI: 88.7, 99.9) and 95.9% (95% CI: 86.0, 99.5) for the two-stages

**Conclusions:** Our questionnaire is valid for the two tested languages and is usable for community-based epilepsy surveys in Ghana. The questionnaire can be adapted for other resource-poor settings, although there will need to be translation and iterative in-country testing to ensure its validity is maintained.

## 1. Introduction

Epilepsy is one of the most prevalent neurological diseases, affecting over 50 million people of all ages globally [1, 2]. Estimates suggest that about 80% of people with epilepsy live in a resource-poor setting with a treatment gap of about 80% [3]. The burden of the disease is disproportionately weighted towards Africa, where more than 10 million people live with epilepsy[4, 5]. In Ghana, the exact prevalence of epilepsy is unknown, but a previous estimation in a rural area suggested it to be 10 per 1000 or 1% of the population [6]. There are no data on burden in urban and peri-urban areas in Ghana.

Africa suffers from a chronic shortage of human resources for health more than any other region [7], with about 0.043 neurologists per 100,000 inhabitants [8]. This lack of personnel has led to task shifting, where skilled staff support and supervise less experienced healthcare workers to provide care at the primary healthcare level [9]. Screening questionnaires are needed to identify people with epilepsy in primary healthcare facilities.

Several epidemiologic surveys of epilepsy have been performed in Low and Middle-income Countries (LMICs) to estimate the prevalence [10]. These surveys often adopt a two-stage study design consisting of a screening phase during which trained field staff interview the population under investigation through a face-to-face meeting with participants. In the subsequent second phase, specialists clinically evaluate positive subjects to confirm true positives [11, 12]. The choice of screening questions is essential, and to validate an instrument is mandatory to establish a screening test’s potential accuracy or inaccuracy. Questions targeting symptoms are vital to maximizing sensitivity (i.e., the proportion of actual cases in the population that test or screen positive), especially in a resource-poor setting with limited access to healthcare. This is important as systematically acquiring clinical information significantly increases the performance of the questionnaire, especially when electroencephalography is unavailable [12].

In epilepsy care and management, antiseizure medication and further interventions depend on the type of seizure, epilepsy syndrome, comorbidity, and other considerations [13]. Solving the problem of epilepsy in resource-poor settings starts with the identification of people with the disease. To this end, developing reliable and accessible screening tools has gained attention [14]. Primary or non-specialist healthcare workers may use an effective screening tool to identify individuals requiring intervention. These screening questionnaires should be in the respondents’ native language and culturally appropriate.

Translating an instrument to the target language must follow pre-determined guidelines [15]. Establishing a tool’s sensitivity, specificity, and predictive value is essential and emphasizes the need for earlier validation [16]. Healthcare providers at the primary and secondary levels of care face many challenges without clinical algorithms or pragmatic rapid diagnostic tests [17]. There may be the need to enforce the widespread use of epilepsy screening questionnaires in primary and secondary healthcare settings to obtain critical information for clinical management and care as a witness. Information from relatives may be inaccurate [18].

We developed an epilepsy screening tool for household surveys. We sought to determine if the tool would improve the identification of all forms of epilepsy in selected health facilities in the Greater Accra Region of Ghana.

## 2. METHODS

### 2.1 Study Population

Our population included children and adults consecutively enrolled from the Shai-Osudoku District Hospital, Prampram Polyclinic, Ningo Health Centre, and the Korle-Bu Teaching Hospital. The cases were people coming for epilepsy treatment with a neurologist or with a physician-confirmed epilepsy diagnosis. Active epilepsy was operationally defined as “two or more unprovoked epileptic seizures separated by at least 24 hours within the last year” [19]. Controls were people attending the same hospitals for other purposes or healthy relatives who had never had a seizure. A neurologist assessed controls to ensure they had no history of seizures. Respondents with an affirmative response to screening questions were deemed screen-positive except if they had febrile convulsions. Cases and controls were matched for geographical location and ethnicity. We excluded those who were severely ill and on admission.

### 2.2 Study design

A cross-sectional study was conducted in the Greater Accra region between August and October 2022 at the outpatient department of selected facilities in the Shai-Osudoku and Ningo-Prampram districts and the Korle-Bu Teaching Hospital. The study is part of the Epilepsy Pathway Innovation in Africa (EPInA) project. (https://epina.web.ox.ac.uk/home).

### 2.3 Epilepsy screening tool development

We conducted a review of evidence on screening tools (questionnaires) previously used in LMICs [16, 20–22] to develop our instrument A local neurologist reviewed the initial version to arrive at the final draft. We then translated and back-translated the tool into the two targeted languages Twi and Dangme. We included questions on convulsive and non-convulsive seizures to cover as many seizure types as possible. We divided the questionnaire into two parts. The first part is administered to either the household head or an adult member with a good knowledge of household members, and stage 2 is administered to people positive in stage 1 (Table 1). For this validation, the seventeen questions were administered to the cases and controls to gauge the sensitivity and specificity of the screening questions.

**Table 1:**
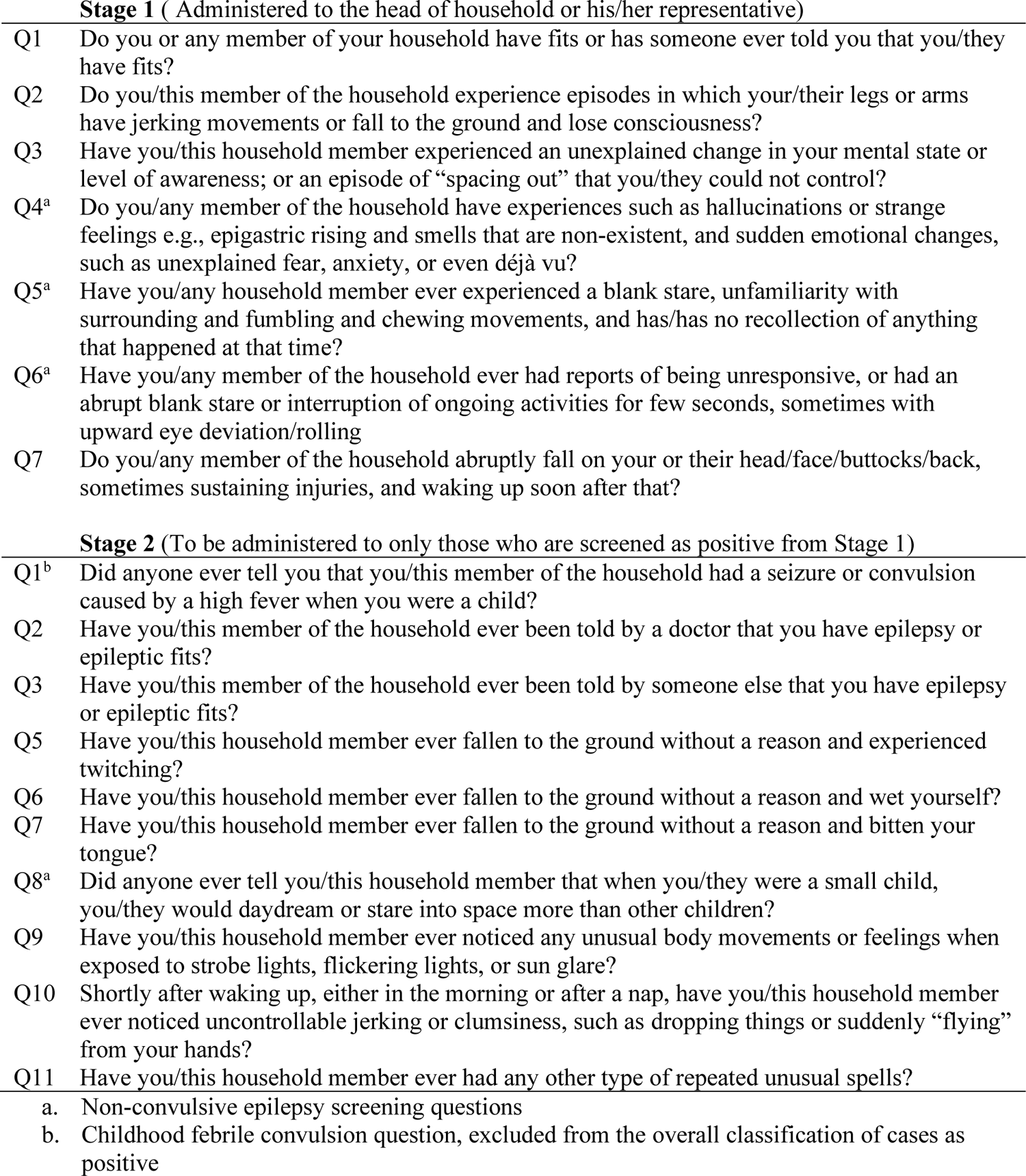
Questionnaire used for screening.

### 2.3 Translation of the screening questionnaire to the local language

Bilingual experts from the Bureau of Ghanaian Languages were consulted to translate the questionnaire into two widely spoken languages in the study setting (Greater Accra region of Ghana): Twi and Dangme, which was back-translated by individuals fluent in the languages who were not involved in the initial translation, following the published guidance on the translation of questionnaires to ensure the meaning remained the same [23]. Jevetic Educational Services did the back translations. Two content experts, a local physician and a neurologist, reviewed the translated versions of the questionnaire to resolve any discrepancies before arriving at the final draft. The questionnaires were also extensively discussed at stakeholder meetings and during training of the research assistants for comprehension, socio-cultural acceptability and adaptability before they were agreed on for the final version.

### 2.4 Sensitivity and Specificity of the Screening Questionnaire

We measured the sensitivity and specificity of the questionnaire by administering the questionnaire to people with confirmed epilepsy and comparing the answers to those without epilepsy. The gold standard for a case was diagnosis by a physician/neurologist with epilepsy expertise. Four trained field workers read the questionnaires verbatim to the participants and recorded the responses. They were, however, not blinded to the diagnosis.

### 2.5 . Sample Size Determination

The sample size was estimated using the formula for testing sensitivity (or specificity) of a single diagnostic test [24] (See supplement 3), which suggested that the questionnaire should be administered to at least 44 people in each language group to have 80% power with 95% confidence to measure the sensitivity and specificity.

### 2.6 Study variables

The questionnaires collected demographic data on age, sex, education, religion, ethnicity, marital status, and the screening questions. The screening questions used at the two stages of the study are shown in Table 1.

### 2.7 Data Management and Statistical Analysis

The data collected on paper questionnaires was entered into Microsoft Excel (version 365) for validation, quality checks and cleaning, after which it was imported into Stata statistical analysis software (version 16)[25] for analysis. Frequencies and percentages were used to describe the demographic data. The Stata module, ‘diagt’ was used to report summary statistics for the diagnostic tests comparing the results from the questionnaire to true disease status.

### 2.8 Classification of cases and controls from screening

For these classifications, if a respondent answered “Yes” to any of the questions used at stages 1 and 2 of the screening (see Table 1), they were classified as positive for that stage, except for Question 1 from Stage 2, which is a question on febrile seizures/convulsions and was not included in such classifications.

### 2.9 Validation of the screening questionnaire

Sensitivity was defined as the proportion of people with the condition correctly identified, and specificity as the proportion of healthy individuals correctly identified. The ROC (Receiver Operating Characteristic curve) defined the area under the curve (for a simple test) the average of sensitivity and specificity.The positive and negative predictive values (PPV & NPV) show the individual’s probability of having the disease following a positive or a negative test. If no prevalence figure is given, the sample is assumed to be a cohort, and PPV & NPV are the proportions of test positives and test negatives identified. Otherwise, they are estimated using the likelihood ratios, assuming the prevalence is correct.

The likelihood ratio of a positive test (LR+) is the ratio of the probability (likelihood) of a positive test result in an affected and in an unaffected individual = Sensitivity/ (1-specificity). Multiplying the prior odds of disease by LR+ gives the odds of disease following a positive test. The likelihood ratio of a negative test (LR-) works similarly. The positive LR is the probability of a positive outcome having a positive screening divided by the likelihood of a negative outcome having a positive screening. The negative LR is the probability of a positive outcome with a negative screening divided by the possibility of a negative outcome having a negative screening. We assessed sensitivity and false-positive rates for the instrument by administering it to individuals with medical record–documented epilepsy or isolated unprovoked seizure and individuals who were seizure-free on medical record review from selected health facilities in two districts in the Shai-Osudoku and Ningo-Prampram districts of the Greater Accra Region of Ghana.

### 2.9A Ethical Considerations

Ethical approval for this study was obtained from the Institutional Review Board of the Dodowa Health Research Centre and the Ghana Health Service Ethics Review Committee, with the following approval numbers: DHRCIRB/95/08/20 and GHS-ERC 022/08/20, respectively. Informed consent was obtained from all study participants. For participants under the age of 18, permission was obtained from one parent or guardian, along with assent from the participant, before their involvement in the study. Participants aged 18 and over provided written informed consent.

During the consenting process, the privacy of the respondents was ensured by conducting study activities in a secluded area with minimal exposure to other individuals. The objectives, justification, and methodologies of the study, as well as the potential risks and benefits, were thoroughly explained to all participants, who were allowed to ask questions. Participation in the study was entirely voluntary. Although immediate or direct benefits were not provided to participants, it was emphasized that their contributions would aid in advancing our understanding of epilepsy and assist in the development of a standardized screening tool for identifying individuals with epilepsy.

Participants were informed of their right to withdraw from the study at any time without facing any repercussions. Responses to screening questions were solicited in a yes or no format. Subsequently, interviews were recorded in Microsoft Excel and transferred to Stata for analysis. All hard and soft data were securely stored in locked file cabinets, with access restricted to the study team. Study participants provided written informed consent.

## 3.0 RESULTS

### Background characteristics of respondents

One hundred and forty participants (70 cases and 70 controls) were recruited for the Dangme questionnaire (Table 2). Among the controls, 40% were young adolescents (age range of 10-19 years); 40% were females, and 34.3% had no formal education (Table 2). Among the cases for the Dangme version, 34.3% were aged 30-39 years; 72.9% were females, and 41.4% had completed Junior High School (JHS)/middle level of formal education (Table 2). The Asante Twi version recruited 100 people (50 cases and 50 controls). Among the 50 controls, 36% were young adults between the age range of 20-29 years, 20% were females, and 26% had completed JHS/Middle school. Among the cases for the Twi version, 24% were between the ages of 30 and 39 years, 60% were females, and 22% had completed some tertiary level of education.

**Table 2:** Background characteristics of respondents.

|  | Dangme |  |  |  | Twi |  |  |  |
| --- | --- | --- | --- | --- | --- | --- | --- | --- |
|  | Cases [N=70] |  | Controls [N=70] |  | Cases [N=50] |  | Controls [N=50] |  |
|  | N | % (95% CI) | N | % (95% CI) | N | % (95% CI) | N | % (95% CI) |
| Age (years) |  |  |  |  |  |  |  |  |
| <10 | 0 | - | 6 | 8.6 (3.8, 18.0) | 0 | - | 2 | 4.0 (1.0, 15.1) |
| 10-19 | 2 | 2.9 (0.7, 11.0) | 28 | 40.0 (29.1, 52.0) | 1 | 2.0 (0.3, 13.4) | 13 | 26.0 (15.5, 40.2) |
| 20-29 | 7 | 10.0 (4.8, 19.7) | 13 | 18.6 (11.0, 29.6) | 7 | 14.0 (6.7, 27.0) | 18 | 36.0 (23.7, 50.4) |
| 30-39 | 24 | 34.3 (24.0, 46.3) | 15 | 21.4 (13.2, 32.8) | 12 | 24.0 (14.0, 38.1) | 9 | 18.0 (9.5, 31.5) |
| 40-49 | 23 | 32.9 (22.8, 44.8) | 5 | 7.1 (3.0, 16.3) | 7 | 14.0 (6.7, 27.0) | 4 | 8.0 (3.0, 19.9) |
| 50-59 | 6 | 8.6 (3.8, 18.0) | 2 | 2.9 (0.7, 11.0) | 12 | 24.0 (14.0, 38.1) | 1 | 2.0 (0.3, 13.4) |
| 60+ | 8 | 11.4 (5.7, 21.4) | 1 | 1.4 (0.2, 9.8) | 11 | 22.0 (12.4, 35.9) | 3 | 6.0 (1.9, 17.4) |
| Sex |  |  |  |  |  |  |  |  |
| Female | 51 | 72.9 (61.1, 82.1) | 28 | 40.0 (29.1, 52.0) | 30 | 60.0 (45.6, 72.8) | 10 | 20.0 (10.9, 33.7) |
| Male | 19 | 27.1 (17.9, 38.9) | 42 | 60.0 (48.0, 70.9) | 20 | 40.0 (27.2, 54.4) | 40 | 80.0 (66.3, 89.1) |
| Education |  |  |  |  |  |  |  |  |
| None | 13 | 18.6 (11.0, 29.6) | 24 | 34.3 (24.0, 46.3) | 2 | 4.0 (1.0, 15.1) | 4 | 8.0 (3.0, 19.9) |
| Primary | 13 | 18.6 (11.0, 29.6) | 21 | 30.0 (20.3, 41.9) | 4 | 8.0 (3.0, 19.9) | 10 | 20.0 (10.9, 33.7) |
| JHS/Middle School | 29 | 41.4 (30.4, 53.4) | 20 | 28.6 (19.1, 40.4) | 17 | 34.0 (22.0, 48.4) | 13 | 26.0 (15.5, 40.2) |
| SHS/O&A Level | 12 | 17.1 (9.9, 28.0) | 4 | 5.7 (2.1, 14.5) | 5 | 10.0 (4.1, 22.3) | 14 | 28.0 (17.1, 42.3) |
| Tertiary | 3 | 4.3 (1.4, 12.7) | 1 | 1.4 (0.2, 9.8) | 22 | 44.0 (30.7, 58.2) | 9 | 18.0 (9.5, 31.5) |
| Religion |  |  |  |  |  |  |  |  |
| Christian | 66 | 94.3 (85.5, 97.9) | 67 | 95.7 (87.3, 98.6) | 47 | 94.0 (82.6, 98.1) | 47 | 94.0 (82.6, 98.1) |
| Other | 4 | 5.7 (2.1, 14.5) | 3 | 4.3 (1.4, 12.7) | 3 | 6.0 (1.9, 17.4) | 3 | 6.0 (1.9, 17.4) |
| Ethnicity |  |  |  |  |  |  |  |  |
| Akan | 5 | 7.1 (3.0, 16.3) | 5 | 7.1 (3.0, 16.3) | 22 | 44.0 (30.7, 58.2) | 21 | 42.0 (28.9, 56.3) |
| Ewe | 2 | 2.9 (0.7, 11.0) | 2 | 2.9 (0.7, 11.0) | 9 | 18.0 (9.5, 31.5) | 8 | 16.0 (8.1, 29.3) |
| Ga-Dangbe | 58 | 82.9 (72.0, 90.1) | 60 | 85.7 (75.2, 92.2) | 17 | 34.0 (22.0, 48.4) | 18 | 36.0 (23.7, 50.4) |
| Other | 5 | 7.1 (3.0, 16.3) | 3 | 4.3 (1.4, 12.7) | 2 | 4.0 (1.0, 15.1) | 3 | 6.0 (1.9, 17.4) |
| Marital status |  |  |  |  |  |  |  |  |
| Single/Underage | 14 | 20.0 (12.1, 31.2) | 59 | 84.3 (73.6, 91.2) | 10 | 20.0 (10.9, 33.7) | 40 | 80.0 (66.3, 89.1) |
| Married | 47 | 67.1 (55.2, 77.2) | 7 | 10.0 (4.8, 19.7) | 30 | 60.0 (45.6, 72.8) | 8 | 16.0 (8.1, 29.3) |
| Divorced/Separated/ Widowed | 9 | 12.9 (6.7, 23.1) | 4 | 5.7 (2.1, 14.5) | 10 | 20.0 (10.9, 33.7) | 2 | 4.0 (1.0, 15.1) |

Table 3 provides the classification of cases and controls from screening. Seventy (100%) with confirmed epilepsy screened positive on all items at stage I, and only one confirmed epilepsy case screened negative on all item questions at stage II. In comparison, 14 (20%) and 10 (14.28%) controls screened positive at stage I and II in the Dangme questionnaire. One (2%) with confirmed epilepsy screened negative on all items at stage I, and 2 (4%) with confirmed epilepsy screened negative on all item questions at stage 2. In comparison, among the controls, 4 (8%) and 3 (6%) screened positive on all item questions at stage 1 and II in the Asante Twi questionnaire.

**Table 3:** Classification of cases and controls from screening.

|  | Dangme (questionnaire) |  |  | Twi (questionnaire) |  |  |
| --- | --- | --- | --- | --- | --- | --- |
|  | +ve | -ve | Total | +ve | -ve | Total |
| Stage 1 |  |  |  |  |  |  |
| True +ve | 70 | 0 | 70 | 49 | 1 | 50 |
| True -ve | 14 | 56 | 70 | 4 | 46 | 50 |
| Total | 84 | 56 | 140 | 53 | 47 | 100 |
| Stage 2 |  |  |  |  |  |  |
| True +ve | 69 | 1 | 70 | 48 | 2 | 50 |
| True -ve | 10 | 60 | 70 | 3 | 47 | 50 |
| Total | 79 | 61 | 140 | 51 | 49 | 100 |

Sensitivities, specificities, PPVs and NPVs of the overall questionnaire are shown in Table 4. The overall predictive ability of both questionnaires was good. The overall performance was 100% sensitivity and specificity 80%, for the Dangme questionnaire at stage I and 98.6% and 85.7% at stage II. The overall PPV and NPV at stage I was 83.3% and 100% and 87.3% and 98.4% at stage II. The Twi version’s overall sensitivity was 98%, and specificity was 92% at stage I. At stage II the sensitivity was 96% and specificity was 94%. The PPV and NPV were above 90%, and the same for stage II of the questionnaire.

**Table 4:** Validation of the overall classifications from the questionnaire compared to true disease status.

|  | Dangme |  |  | Twi |  |  |
| --- | --- | --- | --- | --- | --- | --- |
|  | Test<br>value | 95% CI |  | Test<br>value | 95% CI |  |
|  |  | Lower | Upper |  | Lower | Upper |
| Stage 1 |  |  |  |  |  |  |
| Prevalence (%) | 50.0 | 41.0 | 58.6 | 50.0 | 40.0 | 60.2 |
| Sensitivity (%) | 100.0 | 88.6 | 94.9 | 98.0 | 89.4 | 99.9 |
| Specificity (%) | 80.0 | 68.7 | 88.6 | 92.0 | 80.8 | 97.8 |
| Positive predictive value (%) | 83.3 | 73.6 | 90.6 | 92.5 | 81.8 | 97.9 |
| Negative predictive value (%) | 100.0 | 93.6 | 100.0 | 97.9 | 88.7 | 99.9 |
| ROC area | 0.90 | 0.85 | 0.95 | 0.95 | 0.91 | 0.99 |
| Likelihood ratio (+) | 5.00 | 3.13 | 7.99 | 12.30 | 4.78 | 31.40 |
| Likelihood ratio (-) | 0.00 | - | - | 0.02 | 0.00 | 0.15 |
| Stage 2 |  |  |  |  |  |  |
| Prevalence (%) | 50.0 | 41.0 | 58.6 | 50.0 | 40.0 | 60.2 |
| Sensitivity (%) | 98.6 | 92.3 | 100.0 | 96.0 | 86.3 | 99.5 |
| Specificity (%) | 85.7 | 75.3 | 92.9 | 94.0 | 83.5 | 98.7 |
| Positive predictive value (%) | 87.3 | 78.0 | 93.8 | 94.1 | 83.8 | 98.8 |
| Negative predictive value (%) | 98.4 | 91.2 | 100.0 | 95.9 | 86.0 | 99.5 |
| ROC area | 0.92 | 0.88 | 0.97 | 0.95 | 0.91 | 0.99 |
| Likelihood ratio (+) | 6.90 | 3.88 | 12.30 | 16.00 | 5.33 | 48.00 |
| Likelihood ratio (-) | 0.02 | 0.00 | 0.12 | 0.04 | 0.01 | 0.17 |

Individual questions’ sensitivity, specificity, PPVs, and NPVs are shown in Table 5. For the Dangme version at stage I, the questions Q1-Q7 had a good sensitivity, while Q1, Q2, Q5, and Q7 had the best sensitivity overall. The sensitivity ranged between 91.5% and 97.1%. The specificity was between 91.4% and 95.7% for all individual questions. With the questions at stage II, the sensitivity was lower with some individual questions compared to the stage I individual questions with most questions having a sensitivity of 60% and below, apart from Q2, Q3, and Q5 with 94.3%, 85.7%, and 94.3%. For the Twi version, all the questions had good sensitivity except Q5, which was 38% at stage I. The specificity was between 96% and 100% for all individual questions. With the questions at stage II, the sensitivity was lower in the Twi version, with most questions having a sensitivity of 54% and below, apart from questions Q2 and Q5 with 84% and 82%. The specificity was between 96% and 100%.

**Table 5:**
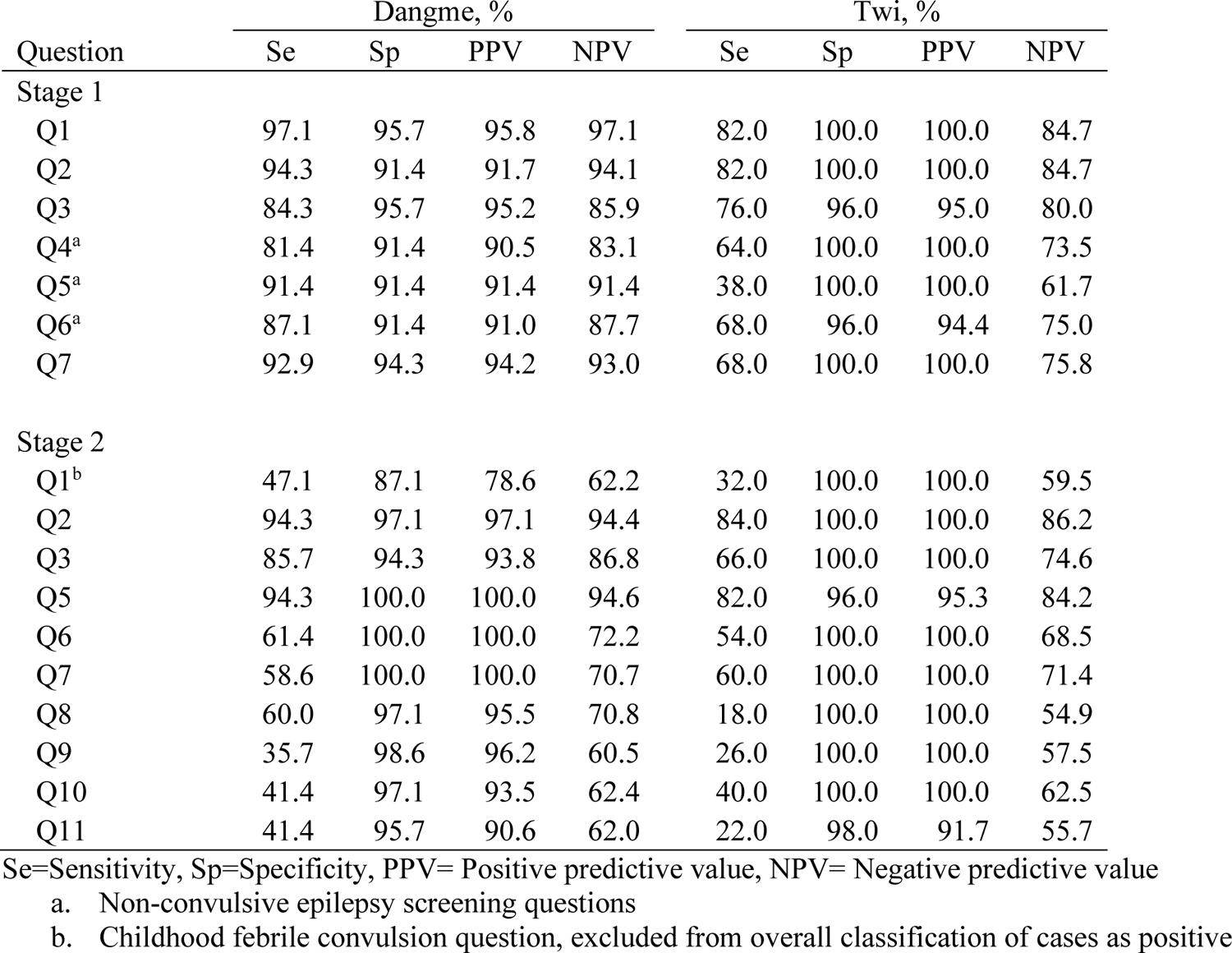
Validation of the individual screening questions compared to true disease status.

| Question | Dangme, % |  |  |  | Twi, % |  |  |  |
| --- | --- | --- | --- | --- | --- | --- | --- | --- |
|  | Se | Sp | PPV | NPV | Se | Sp | PPV | NPV |
| Stage 1 |  |  |  |  |  |  |  |  |
| Q1 | 97.1 | 95.7 | 95.8 | 97.1 | 82.0 | 100.0 | 100.0 | 84.7 |
| Q2 | 94.3 | 91.4 | 91.7 | 94.1 | 82.0 | 100.0 | 100.0 | 84.7 |
| Q3 | 84.3 | 95.7 | 95.2 | 85.9 | 76.0 | 96.0 | 95.0 | 80.0 |
| Q4<sup>a</sup> | 81.4 | 91.4 | 90.5 | 83.1 | 64.0 | 100.0 | 100.0 | 73.5 |
| Q5<sup>a</sup> | 91.4 | 91.4 | 91.4 | 91.4 | 38.0 | 100.0 | 100.0 | 61.7 |
| Q6<sup>a</sup> | 87.1 | 91.4 | 91.0 | 87.7 | 68.0 | 96.0 | 94.4 | 75.0 |
| Q7 | 92.9 | 94.3 | 94.2 | 93.0 | 68.0 | 100.0 | 100.0 | 75.8 |
| Stage 2 |  |  |  |  |  |  |  |  |
| Q1<sup>b</sup> | 47.1 | 87.1 | 78.6 | 62.2 | 32.0 | 100.0 | 100.0 | 59.5 |
| Q2 | 94.3 | 97.1 | 97.1 | 94.4 | 84.0 | 100.0 | 100.0 | 86.2 |
| Q3 | 85.7 | 94.3 | 93.8 | 86.8 | 66.0 | 100.0 | 100.0 | 74.6 |
| Q5 | 94.3 | 100.0 | 100.0 | 94.6 | 82.0 | 96.0 | 95.3 | 84.2 |
| Q6 | 61.4 | 100.0 | 100.0 | 72.2 | 54.0 | 100.0 | 100.0 | 68.5 |
| Q7 | 58.6 | 100.0 | 100.0 | 70.7 | 60.0 | 100.0 | 100.0 | 71.4 |
| Q8 | 60.0 | 97.1 | 95.5 | 70.8 | 18.0 | 100.0 | 100.0 | 54.9 |
| Q9 | 35.7 | 98.6 | 96.2 | 60.5 | 26.0 | 100.0 | 100.0 | 57.5 |
| Q10 | 41.4 | 97.1 | 93.5 | 62.4 | 40.0 | 100.0 | 100.0 | 62.5 |
| Q11 | 41.4 | 95.7 | 90.6 | 62.0 | 22.0 | 98.0 | 91.7 | 55.7 |

Se=Sensitivity, Sp=Specificity, PPV= Positive predictive value, NPV= Negative predictive value
a. Non-convulsive epilepsy screening questions
b. Childhood febrile convulsion question, excluded from overall classification of cases as positive

## 4.0 DISCUSSION

We translated and validated an epilepsy screening questionnaire into two Ghanaian languages, which showed good sensitivity, specificity, PPV, and NPV. We explored the differences in individuals’ sensitivities and specificities and the overall component items of the questionnaire. The likelihood ratios indicated a good but varied probability of predicting epilepsy. These screening tools are simple and can be easily used by health workers at the community level, suggesting a robust epilepsy screening tool for Ghana in these languages. There was limited difference in the specificity of individual and overall item questions, but we noted significant differences in the sensitivity across some individual questions (Tables 4 and 5). Specificity measures were also consistent between questions and languages. This means that a positive screen was consistent with a diagnosis of epilepsy”.

We conducted a hospital-based validation of confirmed epilepsy cases compared to controls. It is known that the diagnostic accuracy of such a study design is usually inflated. Cases with more severe forms of the condition and greater awareness of their state of health have a higher chance of being included. These people will likely screen positive with this instrument, possibly leading to overestimating the sensitivity [26]. Consequently, there is a risk of obtaining inaccurate estimates of the disease prevalence when applying a hospital-based validated questionnaire to the general population. The advantage of validation, however, is that the prevalence can be adjusted to the sensitivity and specificity of the screening test [27].

We also noticed some variations in the accuracy measures between the two languages. This might be due to differences in population characteristics, methods of administration of the questionnaires between the various facilities, linguistic features, and translation dynamics [22, 28]. The differences could be attributed to variations in the general literacy and epilepsy awareness and stigma between the two populations. Health facility-based validation studies are not usually as influenced by stigma-related concealment as community- or population-based studies [29]. The differences in sensitivity of some of the individual item questions may also relate to the frequency of the phenomena (e.g., incontinence, absence seizures, jerking movements or falling to the ground). Absence seizures or non-convulsive seizures may often be overlooked by people living with the condition. Low sensitivity of non-convulsive seizure detection has been previously reported [30].

Currently, no comparable studies are available in Ghana using similar screening questionnaires. A study in Nigeria using a nine-item questionnaire reported similar sensitivity and specificity with a slightly lower PPV [22]. A study in Ecuador had a low PPV of 18.3% [20]. The Rochester Epidemiology project also had a PPV of 23% [21]. Our study had a relatively higher PPV, indicating more people with epilepsy were likely to screen positive.

An advantage of this screening tool is that it includes questions that address multiple seizure types, unlike other available screening tools that address only convulsive epilepsy. A further benefit of our study is the estimation of likelihood ratios for the two languages. This is crucial because sensitivity and specificity alone may not provide enough information on the probability of diagnosing epilepsy. Likelihood ratios are more appropriate for comparing individual questions [31].

We found no difficulties in training a local person to administer the screening questionnaires in the two local languages or any challenges with comprehension of the questions. This was mainly because the data collectors were natives who understood the test languages and administered them to respondents who spoke and understood the same.

### 5.0 Limitations

This study was conducted within healthcare facilities. This could result in selection bias as questions were administered to people with confirmed epilepsy who could differ from the general population. This selection bias limits the overall generalizability of the results. Our approach could lead to an underestimation of epilepsy at the community level, as less overt cases may be missed. The lack of awareness of epilepsy may result in people failing to seek treatment; these people may not have been captured in our study. Our eventual goal is to validate the tool for use in the general population. Here, information was collected at the facility level and from the cases about their medical histories rather than their relatives. Some individuals might be unaware of specific symptoms, which could explain why some cases screened negative. The use of proxies or caregiver questions was an option we considered for stage I of our screening tool. We did not explore this further as some cases did not involve caregivers, and we wanted to ensure consistency across data capture. Future research on the use of proxies or relatives to respond to on behalf of cases would be helpful to see if this can improve reliability, particularly for people with non-convulsive seizures.

## 6.0 Conclusion

We have validated a screening tool for epilepsy that trained community health workers can use in communities to screen for epilepsy in Ghana. Our tool could be adapted to other sub-Saharan African countries following appropriate validation.

## Author Contributions

EKD, JWS, CN and PA conceived and designed the study. EKD and AG analyzed the data. EKD drafted the manuscript with input from PA, AA, CS, EA, SA, JHC, CN, AS, JWS, ADA, and JEW. All authors approved the final draft.

## Data Availability

All data produced in the present study are available upon reasonable request to the authors

## Acknowledgments

This validation was conducted before the larger Epilepsy Pathway Innovation in Africa (EPInA) household survey. EPInA receives funding from NIHR for this research work. This work was carried out at selected health facilities in Ningo-Prampram, Shai-Osudoku District and Korle-Bu Teaching Hospital. We want to thank the heads of the various facilities and staff for helping with the research work. We acknowledge those who contributed to the translation processes of the questionnaires: the Bureau of Ghana Languages for the forward translations and the Jevetic Education Services for the back translations. We are also grateful to the research assistants Daisy Ampadu, Irene Siaw, Andrews Tettey and Edmund Owusu for their assistance with data collection. We also thank the study participants whose generous contributions and time made this research possible. EKD is a Ph.D. student working on the EPInA project. JHS holds an endowed chair at UCL Great Ormond Street Institute of Child Health; all her research is supported by the National Institute for Health Research Biomedical Research Centre at Great Ormond Street Hospital (NIHR GOSH BRC). JWS is based at the NIHR University College London Hospitals Biomedical Research Centre, which the UK Department of Health sponsors. The UK Epilepsy Society endows his current position, and he receives research support from the Marvin Weil Epilepsy Research Fund and the Christelijke Vereniging voor deVerpleging van Lijders aan Epilepsie, Netherland. PA is a neurologist and consultant at the Korle-Bu Teaching Hospital and the University of Ghana Medical School. He is the lead researcher on the EPInA project in Ghana. CN is the overall principal investigator for the EPInA project.

## Supplement 1: Dangme version of screening questionnaire

**Table 1:** Questionnaire used for screening.

| **Stage 1** ( Administered to the head of household or his/her representative) |  |
| --- | --- |
| Q1 | Do you or any member of your household have fits or has someone ever told you that you/they have fits? |
| Q2 | Do you/this member of the household experience episodes in which your/their legs or arms have jerking movements or fall to the ground and lose consciousness? |
| Q3 | Have you/this household member experienced an unexplained change in your mental state or level of awareness; or an episode of “spacing out” that you/they could not control? |
| Q4<sup>a</sup> | Do you/any member of the household have experiences such as hallucinations or strange feelings e.g., epigastric rising and smells that are non-existent, and sudden emotional changes, such as unexplained fear, anxiety, or even déjà vu? |
| Q5<sup>a</sup> | Have you/any household member ever experienced a blank stare, unfamiliarity with surrounding and fumbling and chewing movements, and has/has no recollection of anything that happened at that time? |
| Q6<sup>a</sup> | Have you/any member of the household ever had reports of being unresponsive, or had an abrupt blank stare or interruption of ongoing activities for few seconds, sometimes with upward eye deviation/rolling |
| Q7 | Do you/any member of the household abruptly fall on your or their head/face/buttocks/back, sometimes sustaining injuries, and waking up soon after that? |
| **Stage 2** (To be administered to only those who are screened as positive from Stage 1) |  |
| Q1<sup>b</sup> | Did anyone ever tell you that you/this member of the household had a seizure or convulsion caused by a high fever when you were a child? |
| Q2 | Have you/this member of the household ever been told by a doctor that you have epilepsy or epileptic fits? |
| Q3 | Have you/this member of the household ever been told by someone else that you have epilepsy or epileptic fits? |
| Q5 | Have you/this household member ever fallen to the ground without a reason and experienced twitching? |
| Q6 | Have you/this household member ever fallen to the ground without a reason and wet yourself? |
| Q7 | Have you/this household member ever fallen to the ground without a reason and bitten your tongue? |
| Q8<sup>a</sup> | Did anyone ever tell you/this household member that when you/they were a small child, you/they would daydream or stare into space more than other children? |
| Q9 | Have you/this household member ever noticed any unusual body movements or feelings when exposed to strobe lights, flickering lights, or sun glare? |
| Q10 | Shortly after waking up, either in the morning or after a nap, have you/this household member ever noticed uncontrollable jerking or clumsiness, such as dropping things or suddenly “flying” from your hands? |
| Q11 | Have you/this household member ever had any other type of repeated unusual spells? |
| a. | Non-convulsive epilepsy screening questions |
| b. | Childhood febrile convulsion question, excluded from the overall classification of cases as positive |

## Supplement 3: The formula for sample size Calculation

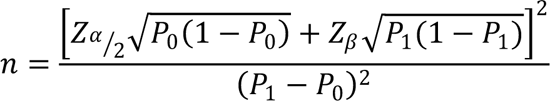

Where *P*_0_is the pre-determined value of sensitivity (or specificity) of the diagnostic questionnaire in English set at 98%, *P*_1_is the expected sensitivity (or specificity) of each of the Asante Twi and Dangme translated questionnaires also set at 90%, *Z*α/2 = 1.96 and *Z*_β_ = 0.84 are critical values from the standard normal distribution for a 95% confidence level and 80% power in estimating differences in diagnostic capabilities of the tools compared to the English one. Using the above figures, the minimum sample size required for each group (case or control) for the Asante Twi and Dangme languages was 44 persons.

## REFERENCES

1. Fiest, K.M., et al., Prevalence and incidence of epilepsy: a systematic review and meta-analysis of international studies. Neurology, 2017. 88(3): p. 296–303.

2. Mbuba, C.K., et al., The epilepsy treatment gap in developing countries: a systematic review of the magnitude, causes, and intervention strategies. Epilepsia, 2008. 49(9): p. 1491–1503.

3. Meyer, A.C.L., et al., Critical determinants of the epilepsy treatment gap: a cross-national analysis in resource-limited settings. Epilepsia, 2012. 53(12): p. 2178–2185.

4. Ba-Diop, A., et al., Epidemiology, causes, and treatment of epilepsy in sub-Saharan Africa. The Lancet Neurology, 2014. 13(10): p. 1029–1044.

5. Prevett, M., Epilepsy in sub-saharan Africa. Practical neurology, 2013. 13(1): p. 14–20.

6. Ae-Ngibise, K.A., et al., Prevalence and risk factors for Active Convulsive Epilepsy in Kintampo, Ghana. Pan Afr Med J, 2015. 21: p. 29.

7. Afriyie, D.O., J. Nyoni, and A. Ahmat, The state of strategic plans for the health workforce in Africa. BMJ global health, 2019. 4(Suppl 9): p. e001115.

8. Kissani, N., et al., Why does Africa have the lowest number of Neurologists and how to cover the Gap? Journal of the Neurological Sciences, 2022. 434: p. 120119.

9. Willcox, M.L., et al., Human resources for primary health care in sub-Saharan Africa: progress or stagnation? Human resources for health, 2015. 13: p. 1–11.

10. Espinosa-Jovel, C., et al., Epidemiological profile of epilepsy in low income populations. Seizure, 2018. 56: p. 67–72.

11. Giuliano, L., et al., Usefulness of a smartphone application for the diagnosis of epilepsy: validation study in high-income and rural low-income countries. Epilepsy & Behavior, 2021. 115: p. 107680.

12. Reutens, D.C., et al., Validation of a questionnaire for clinical seizure diagnosis. Epilepsia, 1992. 33(6): p. 1065–1071.

13. Stern, J.M., Overview of treatment guidelines for epilepsy. Current Treatment Options in Neurology, 2006. 8(4): p. 280–288.

14. Wang, Z., Z. Luo, and S. Li, Anxiety screening tools in people with epilepsy: a systematic review of validated tools. Epilepsy & Behavior, 2019. 99: p. 106392.

15. Cramer, J.A., et al., Development and cross-cultural translations of a 31-item quality of life in epilepsy inventory. Epilepsia, 1998. 39(1): p. 81–88.

16. Keezer, M.R., H.K. Bouma, and C. Wolfson, The diagnostic accuracy of screening questionnaires for the identification of adults with epilepsy: a systematic review. Epilepsia, 2014. 55(11): p. 1772–1780.

17. Yaria, J.O. and A. Ogunniyi, Calibration of the epilepsy questionnaire for use in a low-resource setting. Journal of Environmental and Public Health, 2020. 2020.

18. Mannan, J.B. and U.C. Wieshmann, How accurate are witness descriptions of epileptic seizures? Seizure, 2003. 12(7): p. 444–447.

19. Fisher, R.S., et al., ILAE official report: a practical clinical definition of epilepsy. Epilepsia, 2014. 55(4): p. 475–482.

20. Placencia, M., et al., Validation of a screening questionnaire for the detection of epileptic seizures in epidemiological studies. Brain, 1992. 115(3): p. 783–794.

21. Ottman, R., et al., Validation of a brief screening instrument for the ascertainment of epilepsy. Epilepsia, 2010. 51(2): p. 191–197.

22. Watila, M.M., et al., Translation and validation of an epilepsy-screening questionnaire in three Nigerian languages. Epilepsy & Behavior, 2021. 114: p. 107604.

23. Nieuwenhuijsen, M., Design of exposure questionnaires for epidemiological studies. Occupational and environmental medicine, 2005. 62(4): p. 272–280.

24. Hajian-Tilaki, K., Sample size estimation in diagnostic test studies of biomedical informatics. Journal of biomedical informatics, 2014. 48: p. 193–204.

25. LLC, S., STATA 16. 2019, StataCorp LLC College Station, Texas.

26. Mulherin, S.A. and W.C. Miller, Spectrum bias or spectrum effect? Subgroup variation in diagnostic test evaluation. Annals of internal medicine, 2002. 137(7): p. 598–602.

27. Ngugi, A.K., et al., Prevalence of active convulsive epilepsy in sub-Saharan Africa and associated risk factors: cross-sectional and case-control studies. The Lancet Neurology, 2013. 12(3): p. 253–263.

28. Zavala-Rojas, D., A procedure to prevent differences in translated survey items using SQP. 2014.

29. Ngugi, A.K., et al., The validation of a three-stage screening methodology for detecting active convulsive epilepsy in population-based studies in health and demographic surveillance systems. Emerging themes in epidemiology, 2012. 9: p. 1–8.

30. Placencia, M., et al., A large-scale study of epilepsy in Ecuador: methodological aspects. Neuroepidemiology, 1992. 11(2): p. 74–84.

31. Macaskill, P., et al., Assessing the gain in diagnostic performance when combining two diagnostic tests. Statistics in medicine, 2002. 21(17): p. 2527–2546.

